# *“Syphilis is a normal thing that children are born with…*”: Understanding Barriers and Facilitators to Maternal and Congenital Syphilis Care in Kampala, Uganda

**DOI:** 10.64898/2026.10.22.26366202

**Authors:** Annet A. Onzia, Anna-Beatrice Magoro, Agnes B. Naggirinya, Joshua Beinomugisha, Matthew M. Hamill, Cecilia Akatukwasa, Yukari C. Manabe, Rachel King, Rosalind Parkes-Ratanshi

**Author notes:** Shared last co-authorship (Rachel King, Rosalind Parkes-Ratanshi). **Corresponding author:** Rosalind Parkes-Ratanshi, PhD, Centre for Public Health, Queen’s University Belfast, UK.

## Abstract

**Introduction:** Congenital syphilis (CS) remains a major cause of preventable stillbirths and neonatal deaths globally. This qualitative study explored perceptions and experiences of mothers, partners, providers, and policymakers regarding the barriers and facilitators to implementing antenatal and newborn syphilis testing and treatment in Kampala, Uganda.

**Methods:** We conducted 15 in-depth interviews, four focus group discussions, and 16 key informant interviews between March and May 2022 with pregnant and postnatal women, male partners, health workers, traditional birth attendants, and Ministry of Health policymakers. Data were analyzed using the Consolidated Framework for Implementation Research (CFIR) in NVivo 12 and reported following the COREQ checklist.

**Results:** Participants valued syphilis screening and treatment to protect infant health, particularly when they understood the consequences of untreated infection. Facilitators included respectful counselling, phone-based follow-up, and integration of testing into routine antenatal care. Key barriers included fear of painful injections, stigma and disclosure risk, low male-partner involvement, fragmented information systems, limited infant diagnostics, and medication stockouts. Partner notification was hindered by fears of infidelity accusations and relationship conflict. Notably, caregivers frequently refused treatment for asymptomatic infants due to misconceptions that only visibly ill children required care. Facility opportunities to strengthen the syphilis care cascade were identified such patient tracing systems and strengthened supply chains.

**Conclusions:** Addressing CS requires community education, non-parenteral treatment alternatives, and affordable confirmatory neonatal testing. Stronger supply chains, integrated maternal-infant-partner care pathways, better-equipped health workers, male-engagement strategies, and consistent follow-up are needed to accelerate elimination of mother-to-child syphilis transmission in Uganda and similar settings.

**What is already known on this topic:** Congenital syphilis (CS) remains a major cause of preventable stillbirths and neonatal deaths, but the views of people delivering and receiving syphilis care in Uganda are poorly understood.

**What this study adds:** Participants valued CS screening and treatment to protect infants, and facilitators included respectful counselling, phone follow-up and routine antenatal integration. Barriers included fear of injections, stigma, low male-partner involvement, weak information systems, stockouts of supplies, lack of CS confirmatory tests and caregivers refusing prolonged intravenous treatment for infants without symptoms.

**How this study might affect research, practice or policy:** Programmes need community education, male engagement, patient tracing, stronger supply chains and integrated maternal-infant-partner care. Research should prioritise non-injectable, shorter duration infant treatments and affordable neonatal confirmatory testing.

**Summary:** This qualitative study in Kampala, Uganda explored barriers and facilitators to maternal and congenital syphilis care. Key barriers included fear of painful injections, stigma, low male-partner involvement, and treatment refusal for asymptomatic infants. Facilitators included respectful counselling, phone follow-up, and integrating syphilis testing into routine antenatal care.

## Introduction

Congenital syphilis (CS) is the second leading cause of preventable stillbirth worldwide, after malaria, and remains a major driver of adverse birth outcomes in Sub-Saharan Africa (SSA) [1], despite being preventable through maternal screening and intramuscular penicillin [2]. Uganda integrated same-day dual HIV/syphilis testing and treatment into national antenatal care (ANC) guidelines in 2017. Although 99% of women in Uganda attend some ANC, only 68% complete at least four ANC visits, and only 37% start in the first trimester [3, 4].

Qualitative studies have identified barriers and facilitators along the CS prevention cascade. In the Democratic Republic of Congo and Zambia, key barriers included fragmented health systems, supply shortages, limited provider training, stigma, and low knowledge as key barriers [5]. In Zambia, health worker acceptance and quality assurance were essential for sustaining coverage [6]. A recent Ugandan study found misconceptions about syphilis risk and structural barriers to care-seeking, with village health teams emerging as trusted educational partners [7]. The urban Syphilis Treatment of Partners (STOP) trial in Uganda showed that gender dynamics, fear of infidelity accusations, and domestic violence shaped women’s willingness to notify male partners, resulting in low partner treatment uptake that left women at risk of reinfection and infants at continued risk of CS [8]. Collectively, these studies point to individual-level knowledge gaps and stigma, alongside modifiable health-system constraints such as supply shortages, provider training gaps, and weak partner-notification systems, as persistent barriers to maternal syphilis care.

However, prior research has centered on antenatal screening and treatment, leaving a critical gap in the literature on how CS is recognized, diagnosed, and managed once it occurs. Understanding how affected infants are identified and treated, and what shapes that care, is essential for designing tailored prevention strategies. We therefore conducted a qualitative study nested within the Kampala Syphilis Screening and Treatment (KSST) implementation project, carried out from June 2021 to July 2022 at high-volume public health facilities in Kampala District, Uganda. Guided by the Consolidated Framework for Implementation Research (CFIR) [9], we explored stakeholders’ perceptions, experiences, and the barriers and facilitators affecting CS management along the care pathway.

## Methodology

### Project setting and population

The KSST project was conducted at four public health facilities in Kampala District, Uganda: Kisenyi, Kawaala, and Kiswa Health Centres IV, and Kawempe National Referral Women’s Hospital (details in the companion manuscript). Participants were purposively recruited from women and infants engaged through KSST service-delivery activities.

### Qualitative Study Procedures

We explored perceptions, experiences, and barriers and facilitators related to maternal syphilis and CS. Using purposive sampling across all sites to ensure representation of all five stakeholder groups, we conducted 15 in-depth interviews (IDIs), four focus group discussions (FGDs), and 16 key informant interviews (KIIs) between March and May 2022, all face-to-face and with signed informed consent. IDIs and FGDs involved women treated for syphilis in pregnancy, including those identified as ‘high-risk’ (never tested for syphilis before 36 weeks’ gestation, or testing positive in the third trimester, during labor/delivery, or postnatally), as well as women with adverse pregnancy outcomes, women whose infants were diagnosed or treated for CS, and women who declined treatment. Of seven women with adverse pregnancy outcomes in the main study, three declined participation (**Table 1**). KIIs included health workers, male partners, TBAs, and MOH sexually transmitted infection (STI)/HIV representatives (**Table 2**) and continued until thematic saturation. FGD and IDI findings informed the KIIs.

**Table 1:** Characteristics of participants of Focus Group Discussions and In-depth Interviews.

| Variable |  | Number (%) |
| --- | --- | --- |
| Age | 15-24 | 27 (62.8) |
|  | ≥ 25 | 16 (37.2) |
| Level of education | Informal education | 4 (9.3) |
|  | Primary and below | 24 (55.8) |
|  | Above primary level | 15 (34.9) |
| Marital status | Married | 32 (74.4) |
|  | Single | 3 (7.0) |
|  | Divorced/Separated/Widowed | 8 (18.6) |
| Occupation | Housewife | 11(25.6) |
|  | Small-scale business | 21 (48.8) |
|  | Teacher | 4 (9.3) |
|  | Student | 7 (16.3) |
| Religion | Christian | 26 (60.4) |
|  | Muslim | 12 (28.0) |
|  | Others | 5 (11.6) |

**Table 2:** Characteristics of Key Informant Interview Participants.

| Variable |  | Number (%) |
| --- | --- | --- |
| Age 34 (IQR 21-62) | 21-24 | 4 (25) |
|  | ≥ 25 | 12 (75) |
| Level of education | Informal education | 2 (12.5) |
|  | Primary and below | 3 (18.8) |
|  | Secondary and above | 11 (68.7) |
| Marital status | Married | 10 (62.4) |
|  | Single | 3 (18.8) |
|  | Divorced/Separated/Widowed | 3 (18.8) |
| Occupation | Health care worker/policy maker | 7 (43.8) |
|  | Mechanic | 1(6.3) |
|  | Business | 3 (18.8) |
|  | Traditional Birth Attendant | 2 (12.3) |
|  | Taxi driver /Motorbike cyclist | 3 (18.8) |

Semi-structured guides (**Supplemental files 1,2 and 3**), developed from CFIR domains and national guidelines, covered syphilis-related knowledge, attitudes, practices, barriers and facilitators to care, pregnancy and neonatal outcomes, and male partner engagement. Guides were pilot tested (n=4) and refined through daily debriefs. Trained bilingual research assistants with no prior clinical relationships with participants conducted private 30-60-minute interviews in English or Luganda, to reduce social desirability bias. Interviews were audio-recorded and supplemented with field notes.

### Qualitative Data Analysis

Analysis was guided by the CFIR, using a structured codebook that reflected its domains and constructs alongside emergent themes. To enhance credibility and trustworthiness, we triangulated data collection methods and sources, cross-verifying findings across IDIs, FGDs, and KIIs [10]. Two transcripts were used to familiarize the analyst with the data and develop an initial coding framework, after which open coding was applied. The preliminary framework was reviewed with a second coder and a qualitative analyst, who compared codes against transcript data and reached consensus on code definitions and application. The finalized framework and transcripts were imported into NVivo 12 (QSR International) for data management and coding, and refined codebooks (**Supplemental Files 4 and 5**) developed. Findings are reported according to the Consolidated Criteria for Reporting Qualitative Research (COREQ) checklist [11] (**Supplemental File 6**).

### Patient and Public Involvement Statement

Selected members of the public were involved in the design and conduct of this study. Policymakers, service providers, local leaders and women attending ANC reviewed the registers to ensure the questions were clear, culturally appropriate, and relevant to health concerns which facilitated participant engagement. Findings were disseminated to the Ministry of Health to promote awareness and inform future interventions.

### Ethical approval

The Infectious Diseases Institute Research Ethics Committee (IDI REC 008/2022) and the Uganda National Council for Science and Technology (HS2436ES) approved the study. Participants provided written informed consent, with assurance of voluntary withdrawal and confidentiality, and each received time compensation of UGX 30,000 (_≈_USD 8).

## Results

The analysis identified four main themes.

### Theme 1: Individual and community understanding of syphilis and its consequences

#### Knowledge of Transmission

Understanding syphilis transmission varied. While many participants correctly identified condomless sexual contact and mother-to-child vertical transmission as the primary routes, misconceptions were common.

> *"What I have heard, [it] may be possible through body contact. Let us assume if you wear someone’s clothes, you have urinated, maybe you have gone to a dirty place, you can contract it."* (FGD _Pregnant woman)

#### Recognition of signs and symptoms

Awareness ranged from accurate recognition of skin and genital manifestations to a lack of awareness, as many had asymptomatic infections.

> *"People say it makes your mouth and feet smell, causes wounds, cracks, or itchy private parts."* (KII_Male Partner)

#### Perceived consequences of untreated syphilis and its role in motivating care

Participants consistently described incomplete or delayed syphilis treatment as a reason for adverse outcomes

> *"They gave me medication, but I was not taking it as instructed, and I also didn’t tell my husband, leading to the loss of my pregnancy."* (IDI_non-pregnant woman)

For babies who survive, CS was perceived by some as causing severe, lasting harm, chronic wounds, disability, and congenital anomalies.

> *"She can give birth to a child without some body parts, without an eye or ear; that is the little I know about it"* (KII _Male Partner)

### Theme 2: Facilitators of engagement in care

Participants also identified factors that made testing, treatment, and follow-up more acceptable and feasible (**Figure 1**).

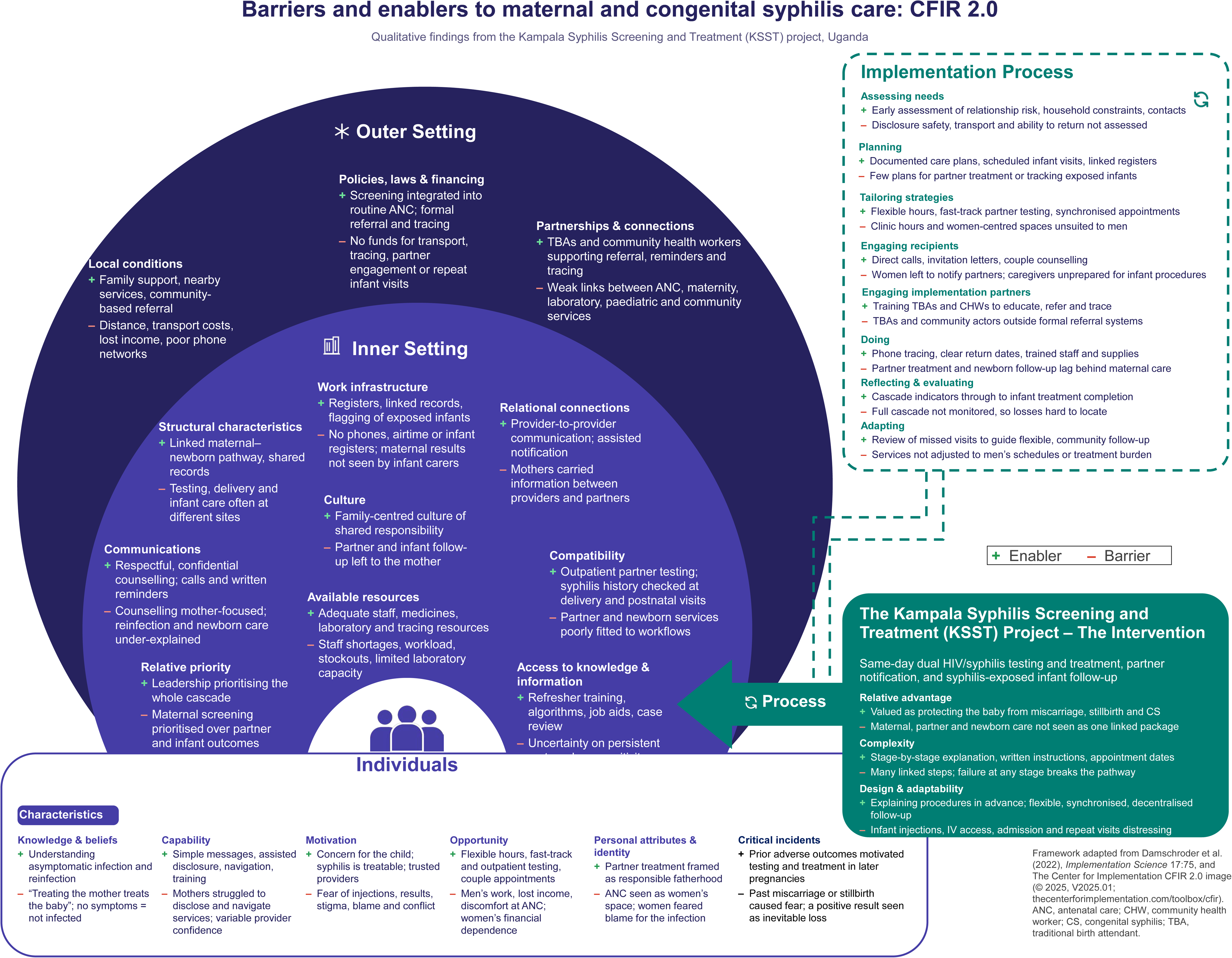

#### Counselling and active communication

Clear, respectful results dissemination and non-judgmental counselling and education reduced fear, restored women’s confidence after diagnosis, and made mothers more willing to start treatment and return with their babies.

> *"I didn’t fear that much because the health worker gave me courage and told me that it’s treatable… from there I became strong."* (IDI_Mother of Treated Baby)

Active outreach, particularly phone contact about results or visits, reduced uncertainty and signaled continued follow-up and care which were described as essential.

> *"This project gave us airtime, so we can now call them to find out if they completed treatment. Also, remind them to bring their husbands."* (KII 1_ Healthcare Worker)

#### Concern for the child as a source of motivation

Across mothers, partners, and caregivers, the baby’s well-being was a powerful motivator: engagement was greater when treatment was framed as protecting the child rather than blaming the mother and allowed mothers to persevere even through painful treatment.

> *"I wish the baby a healthier life because they know nothing about it. I am an adult and not scared of anything. So, I am protecting the baby from the outcomes and danger later on, for example, I have accepted the painful injections."* (FGD _ Pregnant woman)

#### Integration of screening within routine antenatal care

Including maternal screening within routine antenatal services, reduced the need for a separate visit and normalized testing as standard pregnancy care. The first antenatal visit was seen as critical, especially increased home deliveries during the COVID period.

> *"We are lucky that a significant proportion of our mothers still come for antenatal care during this COVID era…that first touchpoint lets us test, counsel, and treat before they leave."* (KII_ Healthcare worker)

### Theme 3: Barriers to syphilis care and management (Figure 1)

#### Subtheme 1: Social and Relational Barriers

##### Therapeutic pluralism: herbs before needles

Many participants initially relied on traditional herbal remedies, often unaware of clinical alternatives, a pattern reinforced by family members.

> *"I didn’t know there were tablets, so when I went back home, my mom told me there was herbal medicine I could take."* (IDI_Pregnant woman)

##### Fear and pain of treatment, including newborn treatment

Experience and fear of painful injections deterred both initiation and completion of treatment.

> *"They used a big syringe like for a cow…When it was time for my next dose, the first shot was still hurting… I withdrew and just watched the syphilis grow."* (IDI_Pregnant woman)

This fear was magnified for newborns: caregivers found repeated injections distressing, sometimes making treatment appear unsafe. Most persisted once they understood why treatment was necessary; refusal was most frequent when that reasoning had not been explained.

> *"I can’t take my baby for those injections. Maybe if they invent another less painful medication, then I will take my child for testing and medication."* (IDI_Pregnant woman)
>
> *“… they had pricked all the veins, and when they had pricked all the hands, I discontinued giving him.”* (IDI_ Breastfeeding woman)

##### Marital stigma, disclosure risk, and fear of relationship conflict

Syphilis was strongly perceived to be associated with infidelity, making disclosure emotionally and socially fraught:

> *“They fear being accused of cheating by their husbands.”* (KII_Healthcare Worker)

Some women delayed disclosure or were treated without telling their partners, increasing the risk of reinfection.

> *“They gave me medication, but I was not taking it as instructed, and I also didn’t tell my husband. I think this led to the loss of my pregnancy.” (IDI_Mother with adverse pregnancy outcome)*

##### Low male-partner engagement and gendered perceptions of antenatal services

Male engagement was among the most difficult parts of the care pathway, as some men saw postnatal clinics as women’s spaces, attendance meant lost work and income.

> *“It is the mother’s responsibility to take the baby for testing like other women do. I must look for money…what will they eat?”* (KII_Male partner)

##### Myths and misconceptions about CS

Some participants believed CS was a normal condition all children are born with, or that a positive test needed no treatment unless the child looked ill; these beliefs reduced perceived need for treatment.

> *"If I can’t see wounds or bad smells on my baby, how can I believe it has syphilis?… I will not allow injections until I see these signs."* (IDI_Breastfeeding woman)
>
> *“Syphilis is a normal thing that children are born with……but syphilis is dangerous in children only in case a child falls sick.”* (IDI_ Pregnant woman)

#### Subtheme 2: Health System and Resource Barriers

##### Fragmentation across services and weak information transfer

This weakened the maternal-newborn pathway, as women often tested, delivered, and received postnatal care at different facilities without shared medical records, so identifying an exposed infant depended on the mother recalling her own diagnosis, and some women were never told to bring their baby for testing.

> *"When getting treated for syphilis, the health workers did not tell me to also test my child."* (IDI_Mother of a newborn who was not tested)

##### Gaps between diagnosis and treatment, and weak follow-up and tracing systems

Even where testing occurred, timely treatment often failed: facilities lacked coordinated systems to ensure completion, and follow-up depended on individual provider initiative rather than a standardized system.

> *"The testing may be done…the kit may be there, but the opportunity for treatment and follow-up to ensure that partners get treated is missing."* (KII_Health care Worker)

##### Barriers to confirmatory infant testing and paternal consent

Confirmatory infant tests were widely described as expensive, rare, or unavailable, leading clinicians to treat presumptively rather than confirm infant infection directly.

> *“Those tests are very expensive…so if the mother is positive, I just treat her.”* (KII_ Healthcare worker)
>
> *“I’m not confident we can test a newborn and say for sure it has congenital syphilis. Those tests are hard to find and cost too much.”* (KII _Policy maker)

Health workers also identified a lack of paternal consent as a significant barrier to infant treatment, particularly where fathers were not involved in antenatal care, reflecting cultural norms that position fathers as key household decisionmakers making their consent a frequent prerequisite for treatment.

> *"It is the father to agree first. Yes, it is consent from the dad. If the dad refuses, the child is not going to get the treatment."* (KII_Healthcare worker)

##### Provider workload, staffing shortages, and limited clinical confidence

Health workers described insufficient time and staff for education, testing, treatment, and follow-up alongside broader duties. Project-supplied extra staff dedicated to syphilis services markedly improved care, though sustainability beyond project support was uncertain.

> *"They gave us more nurses to handle only syphilis services…things are now better…but after this project ends…we don’t know."* (KII_Healthcare Worker)

##### Negative facility experiences

Reports of healthcare workers with judgmental attitudes and long waiting time discouraged women, particularly those who delivered at home from returning for postnatal care.

> *"Some health workers are rude and delay patients, making mothers who gave birth at home too afraid to go back."* (KII_Traditional Birth attendant)

##### Medication stockouts and financial/logistical costs

Stockouts meant women could test positive yet be unable to access treatment on-site, undermining confidence that counselling alone would translate into completed treatment, a gap project-supplied medication directly addressed.

> *"At times, the test is positive, but at the end, no medicine…So, what are you going to do with her? You educate her on her health and counsel her to get the medicine outside…This project at least gives them all the medicines if the test has reacted."* (KII_Healthcare worker)

Informal facility charges prevented some families from completing care even when medication was nominally free, at times pushing women toward traditional providers who lack the training to screen, diagnose, or treat syphilis in pregnancy.

> *"Even if the drugs are free, some health workers still ask for money, so they always come to me instead."* (KII_Traditional birth attendant)

### Theme 4: Opportunities and Recommendations to strengthen the syphilis care cascade

Participants shared recommendations to strengthen the maternal-partner-newborn syphilis care cascade, including expansion of community sensitization and preconception screening, innovations in CS testing and treatment, ensuring a single maternal-infant care pathway with facility-supported follow-up systems, supporting male partners’ testing, and supply chain strengthening (**Table 3**).

**Table 3:** Opportunities and recommendations to strengthen syphilis care.

| Recommendation | Key Elements | Supporting Quotes |
| --- | --- | --- |
| Expansion of community sensitization and preconception screening | Use of radio, TV, and brochures (in local languages) to raise awareness about syphilis.<br>Targeting couples before pregnancy so they know their syphilis status early. | *"Radio, TV, and brochures in English and Luganda could teach people how to prevent and treat syphilis."*_KII_Policy maker<br>*"Don't wait until pregnancy, target those planning to conceive so they test early and know their status beforehand."*_IDI_Pregnant woman |
| Building a single, documented maternal-infant care pathway | Integration of maternal and newborn care into one documented pathway: systematically flag exposed infants, synchronize appointments, provide clear instructions, and review maternal records at delivery and postnatal visits, so newborn follow-up no longer depends solely on a mother recalling her diagnosis. | *"A registry... would ensure mothers, babies, and partners complete treatment."*_KII_Policy maker |
| Supporting partner testing | Expand partner-inclusive education, which providers say accelerates testing and treatment uptake; make couples testing mandatory with joint treatment when both partners test positive; and make partner services more feasible through flexible hours, fast-track testing, and delivery in settings men already use. | *"These days, men escort their wives, making it easy for us to explain the disease and fast-track both partners through care."*_KII_Policy maker |
| Establishing facility-supported tracing systems | Strengthen newborn follow-up through maternal-infant registers, appointment reminders, phone-based tracing, and routine review of maternal records, shifting tracing from individual initiative to a standardized facility system. | *"They called me to remind me to complete the injections...even after I gave birth, again they called to remind me to bring the baby for testing."*_IDI_Breastfeeding woman |
| Innovations in congenital syphilis treatment and testing | Improving medications and creating shorter, less burdensome newborn regimens to reduce distress and improve treatment completion.<br>Developing reliable low-cost confirmatory infant testing | *"Syphilis treatment has remained the same since I was born. If the government could improve the injection or introduce a better formula, more people would complete treatment."*_FGD_Pregnant woman<br>*"We need a simple, low-cost test to confirm congenital syphilis in infants...we are just* |
|  |  | *gambling with things."*_ KII_Policy maker |
| Strengthening policy, supply chains, and reducing household costs | Enforcing existing national protocols requiring treatment of both index cases and partners; leverage the HIV/Syphilis dual rapid test for universal screening at first contact; ensure reliable, decentralized diagnostics and medication; provide transport support for repeated visits. | *"Policies recommend universal syphilis and HIV screening in MCH settings. With the DUO test, we can reach all mothers at first contact."*<br>_KII_Policy maker |

## Discussion

This CFIR-guided qualitative study examined the barriers and facilitators shaping maternal and CS care across four public health facilities in Kampala, Uganda. By engaging stakeholders across the full care pathway, we identified individual, relational, and health-system factors that help explain both the KSST project’s gains and the persistent gaps seen in our companion quantitative analysis.

The facilitators were mapped mainly onto CFIR’s intervention characteristics and process domains. Respectful, non-judgmental counselling, active phone-based follow-up, and integration of syphilis testing into routine antenatal care reduced fear and built trust, helping explain the rise in screening and same-day treatment coverage reported in our companion paper.

Lower coverage was linked to stockouts of rapid test kits and benzathine penicillin and limited laboratory capacity, all within CFIR’s Inner Setting. Similar shortages have constrained syphilis treatment in Zambia and Ghana [12, 13], suggesting these barriers are widely shared.

The barriers most resistant to change clustered within the CFIR Outer Setting (gendered norms, stigma, financial pressures), which typically shift more slowly than individual knowledge or attitudes. Women feared that disclosure would trigger accusations of infidelity, or intimate partner violence, reflecting stigma and unequal gender power dynamics. This left them vulnerable to reinfection from untreated partners. This is consistent with prior research [14–16] and with our earlier STOP study, in which only 18.3% of male partners of syphilis-positive pregnant women received treatment [17]. Men saw antenatal clinics as women’s spaces, and lost income further deterred attendance. Male engagement strategies must therefore address both structural and normative barriers; couples-based testing and treatment within ANC remains a missed opportunity [18–20].

Limited syphilis knowledge likely suppressed demand for screening and treatment [5, 21], and fear of painful benzathine penicillin injections led some women to delay or refuse treatment seen previously in Uganda and Malawi [7, 8, 22].

Our findings help explain the infant treatment attrition in our companion paper, with 46% of infants born to high-risk women met criteria for confirmed or possible CS, yet only 41% completed treatment. Mothers often declined treatment for infants who appeared well, questioning the need for prolonged intravenous therapy or fearing hospitalization. Health workers faced diagnostic uncertainty because confirmatory tests were costly, and lack of paternal consent where fathers had not been engaged antenatally emerged as a previously underreported barrier. As CS management and treatment attrition have received little attention in SSA syphilis research, these findings are a novel contribution.

Task-sharing, increased staffing, and structured follow-up protocols contributed to the improvements participants described, aligning with CFIR’s Teaming and Engaging constructs. Project-availed treatment supplies and phone-based follow-up were particularly valuable during pandemic-related disruptions.

A key strength is the use of CFIR to systematically identify multilevel barriers and facilitators across stakeholder groups spanning the maternal-infant-partner care pathway. The study also offers novel insight into parental refusal of CS treatment. The work has limitations; three of the seven women with adverse pregnancy outcomes declined participation, which may have biased findings toward more favorable accounts and underrepresented the most severe barriers. The sample came from a single urban district, limiting generalizability to rural or other SSA settings.

Our findings point to structural, rather than purely educational, interventions to close the partner and infant testing and treatment gaps, with implications for syphilis management guidelines. A single, documented maternal-infant care pathway, with systematic flagging of exposed infants, synchronized appointments, and dedicated tracing registers, would reduce reliance on mothers self-reporting their diagnosis. Sustaining the KSST gains requires reliable diagnostics and benzathine penicillin supply chains, dedicated staffing or task-shifting, and low-cost tracing resources such as phone airtime. Less painful or shorter treatment regimens with targeted counselling could improve treatment completion, particularly for newborns, and affordable point-of-care confirmatory infant diagnostics are urgently needed to reduce over-treatment and missed CS cases.

## Conclusions

Respectful counselling, phone-based follow-up, and integration of syphilis testing into routine antenatal care helped women and infants engage with treatment, but barriers such as fear of painful injections, stigma, low male-partner involvement, fragmented information systems, limited infant diagnostics, and medication stockouts, along with refusal of treatment for asymptomatic infants, point to the need for community education, non-parenteral alternatives, affordable confirmatory neonatal testing, stronger supply chains, integrated maternal-infant-partner pathways, better-equipped health workers, and consistent follow-up to accelerate elimination of mother-to-child syphilis transmission in Uganda and similar settings.

## Data Availability

The focus group discussion, in-depth interview, and key informant interview guides are attached in the additional files. The source data is in recorded audio, which could break the anonymity of participants. The IDIs and KIIs were anonymized and transcribed. The data analyzed for this manuscript will not be made available online because of its sensitive nature, but the corresponding author can be contacted on a case-by-case basis to provide data following a reasonable request.

## Acknowledgements

The authors express their gratitude to all the study participants and the participating health facilities in Kampala District that allowed the study to be conducted within their premises. We are grateful to all the research assistants for their contribution to collecting data.

## Declarations

### Authors’ Contributions

All authors contributed to the study. AAO: Project administration, data collection, data analysis, draft manuscript writing. AAO is the guarantor; ABM: Data collection, data analysis, manuscript reviews; ABN: Project design and administration, supervision, manuscript reviews; JB: Qualitative data collection, data analysis; YCM: Supervision, manuscript reviews; MMH: Supervision, manuscript reviews; RK: Qualitative analysis, manuscript reviews; CA: Qualitative data analysis, draft manuscript writing and reviews; RPR: Funding acquisition, project design, supervision, manuscript draft and reviews. All authors read and approved the final manuscript.

### Consent for publication

Not Applicable

## List of abbreviations

ANC: Antenatal Care
CFIR: Consolidated Framework for Implementation Research
COREQ: Consolidated Criteria for Reporting Qualitative Research
CS: Congenital Syphilis
FGD: Focus Group Discussion
HIV: Human Immunodeficiency Virus
IDI: In-depth Interview
IDI REC: Infectious Diseases Institute Research Ethics Committee
KII: Key Informant Interview
KSST: Kampala Syphilis Screening and Treatment project
SSA: Sub-Saharan Africa
UNCST: Uganda National Council of Science and Technology
WHO: World Health Organization

## REFERENCES

1. Korenromp, E.L., et al., Correction: Global burden of maternal and congenital syphilis and associated adverse birth outcomes—Estimates for 2016 and progress since 2012. PLoS One, 2019. 14(7): p. e0219613.

2. Organization, W.H., Global health sector strategies on, respectively, HIV, viral hepatitis and sexually transmitted infections for the period 2022-2030. 2022: World Health Organization.

3. Saham, N., et al., Barriers to Routine Antenatal Syphilis Screening in Uganda: Provider Perspectives and Practices. Glob Qual Nurs Res, 2025. 12: p. 23333936251375457.

4. Kazibwe, A., et al., Facilitators, barriers and service availability for delivering integrated care for the triple elimination of HIV, syphilis and hepatitis B vertical transmission in Uganda: a multi-site explanatory mixed methods study. BMC Health Serv Res, 2025. 25(1): p. 626.

5. Nkamba, D., et al., Barriers and facilitators to the implementation of antenatal syphilis screening and treatment for the prevention of congenital syphilis in the Democratic Republic of Congo and Zambia: results of qualitative formative research. BMC Health Serv Res, 2017. 17(1): p. 556.

6. Ansbro É, M., et al., Introduction of Syphilis Point-of-Care Tests, from Pilot Study to National Programme Implementation in Zambia: A Qualitative Study of Healthcare Workers’ Perspectives on Testing, Training and Quality Assurance. PLoS One, 2015. 10(6): p. e0127728.

7. Miller, A.P., et al., “As long as you are married you cannot protect yourself against syphilis”: qualitative exploration of syphilis risk perception and antenatal care seeking among pregnant women in Uganda. BMC Public Health, 2026. 26(1): p. 657.

8. Nakku-Joloba, E., et al., Perspectives on male partner notification and treatment for syphilis among antenatal women and their partners in Kampala and Wakiso districts, Uganda. BMC Infect Dis, 2019. 19(1): p. 124.

9. Kirk, M.A., et al., A systematic review of the use of the Consolidated Framework for Implementation Research. Implementation Science, 2015. 11(1): p. 72.

10. Damschroder, L.J., et al., Fostering implementation of health services research findings into practice: a consolidated framework for advancing implementation science. Implementation Science, 2009. 4(1): p. 50.

11. Tong, A., P. Sainsbury, and J. Craig, Consolidated criteria for reporting qualitative research (COREQ): a 32-item checklist for interviews and focus groups. Int J Qual Health Care, 2007. 19(6): p. 349–57.

12. Dassah, E.T., Y. Adu-Sarkodie, and P. Mayaud, *Rollout of rapid point of care tests for antenatal syphilis screening in Ghana: healthcare provider perspectives and experiences*. BMC Health Services Research, 2018. 18(1): p. 130.

13. Jones, A.V., et al., Benzathine penicillin G stockouts and other barriers to documented syphilis treatment in pregnancy in Zambia. PLoS One, 2024. 19(6): p. e0304576.

14. Díaz-Olavarrieta, C., et al., Maternal syphilis and intimate partner violence in Bolivia: a gender-based analysis of implications for partner notification and universal screening. Sex Transm Dis, 2007. 34(7 Suppl): p. S42–6.

15. Lech, M.M., Non-effective partner notification system: a missed opportunity for the reduction of sexually transmitted infections in sub-Saharan Africa. Med Wieku Rozwoj, 2003. 7(4 Pt 1): p. 503–9.

16. Pule, M.K., et al., Mapping evidence on youth experiences and practices on partner notification for sexually transmitted infections in sub-Saharan Africa: scoping review. Frontiers in Reproductive Health, 2026. **Volume** 8 **-**2026.

17. Parkes-Ratanshi, R., et al., Low male partner attendance after syphilis screening in pregnant women leads to worse birth outcomes: the Syphilis Treatment of Partners (STOP) randomised control trial. Sexual health, 2020. 17(3): p. 214–222.

18. Foglabenchi, L.H., T. Marchant, and H. Stöckl, Beyond ’Go and bring your husband’: a COM-B guided qualitative study on the barriers to male involvement in antenatal care in Bamenda Health District, Cameroon. PLOS Glob Public Health, 2025. 5(5): p. e0002904.

19. Jefferys, L.F., et al., Official invitation letters to promote male partner attendance and couple voluntary HIV counselling and testing in antenatal care: an implementation study in Mbeya Region, Tanzania. Reprod Health, 2015. 12: p. 95.

20. Sakala, D., et al., *Socio-cultural and economic barriers, and facilitators influencing men’s involvement in antenatal care including HIV testing: a qualitative study from urban Blantyre, Malawi*. BMC Public Health, 2021. 21(1): p. 60.

21. Kanyangarara, M., N. Walker, and T. Boerma, Gaps in the implementation of antenatal syphilis detection and treatment in health facilities across sub-Saharan Africa. PLoS One, 2018. 13(6): p. e0198622.

22. Mbura, J.S., H.N. Mgaya, and H.K. Heggenhougen, The use of oral herbal medicine by women attending antenatal clinics in urban and rural Tanga District in Tanzania. East Afr Med J, 1985. 62(8): p. 540–50.

