## Supplemental figure 1 for "*“Syphilis is a normal thing that children are born with…*”: Understanding Barriers and Facilitators to Maternal and Congenital Syphilis Care in Kampala, Uganda"

**Reducing maternal and congenital syphilis in Uganda; a mixed methods observational study.**

In-Depth Interview Guide

**Introduction:**

Hi, my name is ____________ and I want to thank you for joining us today. I am working together with other scientists at the Infectious Diseases Institute and other institutions on this project in Kisenyi Health Centre IV, Kiswa Health Centre IV, Kawaala HC IV and Kawempe National Referral Hospital. We are conducting this research to explore your perceptions, attitudes and barriers to testing and treatment of maternal and congenital syphilis. I would like to say that there are no right or wrong answers in our conversation. We will simply be discussing your views, opinions and experiences on a range of topics. I would like to audio record our conversation as I may not be able to remember all your responses. Please do not be concerned about this: all measures will be taken by the researchers to maintain confidentiality of the interviews and discussions.

Information you tell us will **ONLY** be used for this research project. As we are audio recording the interview, we ask that you refrain from using names or identifying information of yourself or your partner. If at any time during the interview you feel uncomfortable you can ask for a break, refuse to answer any question, and are always free to leave. Do you have any questions before we start?

**Create rapport:** How have you been? How is work? How is the baby?

1. **Knowledge about syphilis**
   - Had you ever heard about syphilis before? (*Probe; how it is spread, signs and symptoms of syphilis, treatment of syphilis, effects of not treating syphilis to mother and child)*

2. **History of syphilis infection**
   - How did you come to know that you have syphilis? (*Probe whether its first time to test)*
   - How many months along were you when you found out that you have syphilis?
   - How did you feel when you found out that you have syphilis?

INFECTIOUS DISEASES INSTITUTE
RESEARCH ETHICS COMMITTEE (IDI-REC)
APPROVED
VALID UNTIL
24 JUN 2023

3. **Treatment**

- We would like you to describe your experiences with the different services that you received regarding syphilis
    - *Probe on testing -how long it took to test and get results*
    - *Probe on treatment – how long after testing did patient receive treatment and how/ how did you feel about the treatment*
    - *Probe on side effects – How did you handle the side effects*

**2: Partner notification**

After finding out that you have syphilis, did you inform your partner (s) about it?

- **If no,**

o Why were you not able to tell your partner about your syphilis results?
o Did you seek the help of the health worker or someone else to inform your partner about your syphilis results? Explain more. *If no, why?*
o If yes, what was his reaction when the health worker told him? (*Probe if he accepted to test, what were results, treatment)*

- If yes,

o Why and how easy was it for you to inform your partner about your syphilis results
o When you told your partner about it, what was his reaction?
o Did he accept to go for testing?
**Probe;**
**if no, why?**
If yes, what were his results?
If positive, did he get treatment for syphilis?

What are the perceived benefits of informing your partner that you have syphilis?

**3: Birth outcomes**

a) When you delivered your child, was it tested for syphilis?
    - If no, why?

INFECTIOUS DISEASES INSTITUTE
RESEARCH ETHICS COMMITTEE (IDI-REC)
APPROVED
VALID UNTIL
24 JUN 2023

- If yes, *Probe; how she felt, what were the results? Probe; for high risk mother ask if child was tested*

b) If the child was positive, how did you feel?

c) Was the baby given treatment?

- If no, why?
- If yes, (*Probe on type of treatment (IM or IV or syrup), side effects, what she did in case of side effects?)*

d) What are the perceived benefits of having a syphilis positive child treated early?

e) What are the perceived challenges of having a syphilis positive child not treated?

**General questions**

4. What did you like about the syphilis services that were offered?

5. What did you not like about the syphilis services that were offered?

6. What challenges did you face ever since you found out that you have syphilis? (*Probe on partner notification, birth outcomes, treatment)*

7. What were the benefits that you achieved from getting syphilis treatment?

8. Lessons learnt; what did you learn during the time that you were receiving treatment for syphilis?
9. Suggestions; Is there anything that you think should be changed or improved among the syphilis services offered at the health facility?

Thank you so much for taking time to participate in this study.

THE END

INFECTIOUS DISEASES INSTITUTE
RESEARCH ETHICS COMMITTEE (IDI-REC)
APPROVED
VALID UNTIL
24 JUN 2023
