## Supplemental file 2 for "*“Syphilis is a normal thing that children are born with…*”: Understanding Barriers and Facilitators to Maternal and Congenital Syphilis Care in Kampala, Uganda"

**Reducing maternal and congenital syphilis in Uganda; a mixed methods observational study.**

Key Informant Guide

**Introduction:**
Hi, my name is _____________ and I want to thank you for joining us today. I am working together with Dr Annet Onzia on this study in Kisenyi Health Centre IV, Kiswa Health Centre IV, Kawaala HC IV and Kawempe National Referral Hospital. We are conducting this research to explore your perceptions, attitudes and barriers to testing and treatment of maternal and congenital syphilis. I would like to say that there are no right or wrong answers in our conversation. We will simply be discussing your views, opinions and experiences on a range of topics. I would like to audio record our conversation as I may not be able to remember all your responses. Please do not be concerned about this: all measures will be taken by the researchers to maintain confidentiality of the interviews and discussions. Information you tell us will **ONLY** be used for this research project. As we are audio recording the interview, we ask that you refrain from using names or identifying information of yourself or your partner. If at any time during the interview you feel uncomfortable you can ask for a break, refuse to answer any question, and are always free to leave. Do you have any questions before we start?

**Create rapport:** How have you been? How is work? Who do you work as at this hospital/ Health center? How long have you worked here?

### PART A: HEALTH CARE WORKERS INVOLVED IN SYPHILLIS MANAGEMENT AT THE HEALTH CARE FACILITIES

1. **Knowledge about syphilis service provision**
    - Tell us what you know about syphilis service provision here at this Health center/ Hospital.
    - Let us talk about refusal or hesitancy to get treatment for syphilis.
    - a) among pregnant women
    - b) their partners
    - c) Babies tested positive for syphillis
    - What are the perceived challenges of syphilis positive mothers in disclosing to their partners?

2. **Experiences with syphilis infection**
    - How have you been involved in syphilis related service provision? (*probe what the person does, how often)*
    - Tell us about your experiences working with syphilis related service provision? (*Probe on activities-testing, treatment, side effects, follow up, policy and guidelines strengths and weaknesses)*



INFECTIOUS DISEASES INSTITUTE RESEARCH ETHICS COMMITTEE (IDI-REC)
APPROVED
VALID UNTIL
24 JUN 2023

- What beliefs do you think patients might have about syphilis or treatment of syphilis? *(probe about myths and misconceptions regarding treatment)*

3. **What are the mother's perceived Likes and dislikes about syphilis service provision?**
    - Health worker-patient relationship
    - Treatment of congenital syphilis *(Probe mothers' attitudes and practices to baby testing and treatment)*
4. **Challenges**
    - What challenges have you met while offering syphilis related services? (*Probe on APN challenges, treatment challenges, health worker challenges)*
    - How can the above challenges be addressed?
    - What are some of the challenges you think patients may experience in seeking for syphilis treatment.
5. **Benefits**
    - Of what benefits are syphilis related services to patients/patients seeking care
6. **Lessons learnt**
    - What have you learnt while offering syphilis related services?
7. **Recommendations**
    - In your opinion what should be done to improve on syphilis related service provision
8. Might you have any questions, comments or recommendations as regards POC/Self-sample collection issues at community pharmacies?

Thank you so much for invaluable contribution to this study.

**PART B: POLICY MAKERS AT THE MINISTRY OF HEALTH IN SYPHILLIS ELIMINATION PROGRAM**

1. As a policy maker in the elimination of congenital syphilis, what do you think about the current syphilis management guidelines (*Probe for their opinions about these guidelines; benefits; challenges; recommendations to improve them*).

2. What is your opinion about testing babies for congenital syphilis? (*Probe for what they like/benefits and why; challenges/obstacles and why; whether they recommend them and why or why not).*

3. What is your perspective as regards improving syphilis testing and treatment for all pregnant women, their partners and babies? *(Probe for their suggestions/recommendations; what pharmacies can do better; what the Ministry of Health can do better; how the kits can be improved).*



INFECTIOUS DISEASES INSTITUTE
RESEARCH ETHICS COMMITTEE (IDI-REC)
APPROVED
VALID UNTIL
24 JUN 2023

4. What is your opinion as regards patients' notifications? What do you make of the assisted partner notification register that this project designed to help enhance partner notification and treatment? (*Probe for benefits; challenges; whether they would recommend their use; how the service can be improved; ensure a copy of the assisted partner notification register has been sent to them to familiarize themselves with)*

5. What is your opinion about such projects like this, that emphasize baby testing, treatment and follow-up? What do you think about the syphilis birth outcomes register and the assisted*(ensure there is a copy of the birth outcomes register they are familiar with)*

7.What is your recommendation for management of syphilis so as to reduce the burden of congenital syphilis in the country?

8. Do you have any questions or concerns to the project team regarding this project and its objectives? Thank you so much for taking part in this study and for you invaluable contribution.

**END**

INFECTIOUS DISEASES INSTITUTE
RESEARCH ETHICS COMMITTEE (IDI-REC)
APPROVED
VALID UNTIL
24 JUN 2023
