## Supplemental file 4 for "*“Syphilis is a normal thing that children are born with…*”: Understanding Barriers and Facilitators to Maternal and Congenital Syphilis Care in Kampala, Uganda"

### Syphilis Pfizer IDIs

#### Codes\\Thematic coding

thematic analysis

| Name | Description | Files | References |
|---|---|---|---|
| Benefits of testing for syphilis | | 1 | 2 |
| Challenges faced since found out are syphilis positive | | 0 | 0 |
| Not getting cured | | 1 | 1 |
| Challenges patients may face looking for syphilis services | | 0 | 0 |
| Delays | | 1 | 4 |
| Not available in all hospitals | | 1 | 1 |
| EXPERIENCES OF WOMEN WHO TEST POSITIVE | Experiences of women who test positive for syphilis during pregnancy | 8 | 12 |
| Benefits achieved from getting treatment | | 2 | 2 |
| Cured | | 4 | 5 |
| Saved my life | | 1 | 1 |

| Name | Description | Files | References |
|---|---|---|---|
| SUGGESTIONS FROM MOTHERS | | 6 | 8 |
| Suggestions for changes or improvements in syphilis services offered at facility | | 1 | 2 |
| Change treatment for babies | | 1 | 1 |
| Conduct syphilis outreaches | | 1 | 1 |
| Create more space at facility | | 1 | 1 |
| Equip private clinics with testing kits and drugs | | 1 | 1 |
| Improve TAT for test results | | 1 | 1 |
| Need for professionals to inject | | 1 | 1 |
| Sensitization | | 4 | 6 |
| Stock drugs and testing kits in hospitals | | 1 | 2 |
| HISTORY OF SYPHILIS INFECTION | | 0 | 0 |
| Experiences with syphilis services received | | 0 | 0 |
| Drugs not available in some facilities | | 1 | 2 |
| Side effects | | 7 | 14 |

| Name | Description | Files | References |
|---|---|---|---|
| Testing | | 9 | 14 |
| Treatment | | 15 | 56 |
| Use of herbal medicine | | 1 | 2 |
| How did you feel when partner tested positive | | 6 | 10 |
| How did you feel when tested positive | | 9 | 13 |
| How did you know you have the infection | | 13 | 23 |
| When tested negative | | 1 | 1 |
| KNOWLEDGE ABOUT SYPHILIS | | 2 | 2 |
| Benefits of testing when pregnant | | 3 | 3 |
| Effects of not treating | | 3 | 4 |
| Death | | 1 | 2 |
| Giving birth to a child with missing body parts | | 1 | 1 |
| High costs incurred | | 1 | 1 |
| Infect partner | | 1 | 1 |

| Name | Description | Files | References |
|---|---|---|---|
| Infect the newborn | | 8 | 11 |
| Miscarriage | | 3 | 4 |
| Myths | | 3 | 3 |
| Signs and symptoms | | 17 | 46 |
| Spread | | 15 | 37 |
| Treatment | | 9 | 22 |
| Lessons learnt during treatment | | 9 | 17 |
| Availability of healthcare workers | | 2 | 2 |
| Community use of herbal medicine | | 1 | 2 |
| Limited information in the community | | 1 | 1 |
| People don’t want to go to facilities | | 2 | 2 |
| Self-protection | | 2 | 2 |
| Test before treating | | 1 | 1 |
| Testing when or when not pregnant | | 1 | 2 |
| To do routine tests | | 5 | 5 |

| Name | Description | Files | References |
|---|---|---|---|
| MALE PARTNER ATTITUDES TOWARDS TESTING & TREATMENT | | 1 | 1 |
| Always busy | | 1 | 1 |
| Fear | | 1 | 1 |
| PARTNER NOTIFICATION | | 9 | 16 |
| Never notified partner | | 0 | 0 |
| Reasons for not notifying | | 1 | 1 |
| Notified partner | | 8 | 12 |
| Partner not tested | | 3 | 5 |
| Partner reaction | | 4 | 4 |
| Partner tested | | 8 | 13 |
| Why notified partner | | 7 | 8 |
| Perceived benefits of informing partner | | 4 | 4 |
| SYPHILIS SERVICES PROVISION | | 0 | 0 |
| Dislikes | | 0 | 0 |

| Name | Description | Files | References |
|---|---|---|---|
| Likes | | 0 | 0 |
| VIEWS ON SYPHILIS RELATED BIRTH OUTCOMES | | 12 | 49 |
| AEs | | 5 | 15 |
| Baby tested negative | | 2 | 2 |
| Baby tested positive | | 6 | 17 |
| Benefits of testing babies | | 1 | 1 |
| Benefits of treating babies | | 3 | 3 |
| Challenges associated with giving birth to a positive baby | | 3 | 8 |
| Danger in not treating syphilis positive baby | | 3 | 3 |
| Why patients refuse treatment | | 1 | 1 |
| Mindset | | 1 | 1 |
