## Supplemental file 5 for "*“Syphilis is a normal thing that children are born with…*”: Understanding Barriers and Facilitators to Maternal and Congenital Syphilis Care in Kampala, Uganda"

### Syphilis Pfizer KIIs

#### Codes

| Name | Description | Files | References |
|---|---|---|---|
| BENEFITS OF SYPHILIS SERVICES | | 4 | 6 |
| Change in patient perception that its treatable | | 2 | 3 |
| Cured | | 2 | 2 |
| Stop the spread | | 1 | 1 |
| Challenges patients experience in seeking syphilis services | | 5 | 18 |
| Delays to receive treatment | | 2 | 4 |
| Drug stock outs | | 1 | 2 |
| Mismanaged from other facilities | | 1 | 2 |
| With partners | | 5 | 10 |
| EXPERIENCES WITH SYPHILIS INFECTION | | 4 | 13 |

| Name | Description | Files | References |
|---|---|---|---|
| Challenges met while offering services | | 0 | 0 |
| Congestion at facility | | 1 | 2 |
| Injections used to be administered elsewhere | | 1 | 1 |
| Lack communication channels with patients | | 1 | 1 |
| Lack of testing services at facility | | 1 | 3 |
| Limited syphilis treatment information | | 1 | 1 |
| Lost to follow up | | 1 | 2 |
| Mothers with multiple partners | | 1 | 3 |
| Myths surrounding syphilis | | 1 | 1 |
| Being a family disease | | 3 | 5 |
| Not curable | | 1 | 1 |

| Name | Description | Files | References |
|---|---|---|---|
| Need for consent to treat babies | | 1 | 1 |
| Non-disclosure | | 1 | 1 |
| Patient mistrust of government facilities | | 1 | 1 |
| Patients not completing treatment | | 1 | 2 |
| Patients not transparent | | 2 | 4 |
| Easier for men to disclose compared to women | | 1 | 3 |
| Poor mothers | | 1 | 1 |
| Results verification | | 1 | 2 |
| Some clients are busy and can’t wait for long | | 1 | 4 |
| Some men prefer getting treatment outside the referral | | 1 | 1 |
| Some patients fear injections | | 2 | 5 |
| Stigma | | 2 | 3 |

| Name | Description | Files | References |
|---|---|---|---|
| Stockouts | | 4 | 9 |
| Unable to test babies | | 2 | 3 |
| Understaffed | | 4 | 10 |
| Effects to mother | | 1 | 2 |
| Involvement in service provision | | 0 | 0 |
| Side effects | | 2 | 4 |
| Solutions to challenges met while offering syphilis services | | 0 | 0 |
| Follow-ups | | 1 | 2 |
| Mothers and treated in community | | 1 | 2 |
| Partners tested and treated elsewhere | | 1 | 1 |
| Refer for testing | | 1 | 1 |
| Symptoms | | 1 | 1 |
| Treatment | | 3 | 6 |

| Name | Description | Files | References |
|---|---|---|---|
| KNOWLEDGE ABOUT SYPHILIS SERVICE PROVISION | | 5 | 26 |
| Services provided | | 5 | 26 |
| Contact tracing and preventive measures | | 3 | 4 |
| Counselling and health education | | 5 | 12 |
| Disclosure | | 1 | 1 |
| Screenings, testing and treatment | | 4 | 9 |
| LESSONS LEARNT WHILE OFFERING SYPHILIS SERVICES | | 4 | 15 |
| Attained counselling experience | | 1 | 1 |
| Disease is treatable | | 2 | 3 |
| Easy to disclose | | 1 | 2 |
| Many people have the disease | | 1 | 1 |
| Patients lack of right information on syphilis | | 2 | 2 |

| Name | Description | Files | References |
|---|---|---|---|
| Patients need to be given time and attention | | 1 | 3 |
| Patients perceptions and myths about the disease | | 1 | 1 |
| Syphilis is a serious disease | | 2 | 2 |
| Mothers perceived dislikes about syphilis services | | 4 | 7 |
| Painful injection | | 4 | 7 |
| Mothers perceived likes about syphilis services | | 2 | 5 |
| Patient beliefs about syphilis | | 0 | 0 |
| Patient health worker relationship | | 1 | 1 |
| QUESTIONS | | 2 | 5 |
| Recommendations to improve on syphilis related service provision | | 6 | 31 |
| Avail an electronic medical records management system | | 1 | 2 |
| Avail baby testing services | | 2 | 2 |

| Name | Description | Files | References |
|---|---|---|---|
| Avail IEC materials for syphilis and sensitization | | 3 | 6 |
| Empower VHTs | | 1 | 1 |
| Facilitate communication | | 1 | 2 |
| Involve IPs in treatment | | 1 | 1 |
| Policy | | 1 | 1 |
| Provide testing kits and drugs | | 3 | 4 |
| Simplify home visits | | 1 | 1 |
| Target mothers before conception | | 1 | 1 |
| Train and recruit more staff | | 3 | 6 |
| Treat both partners | | 1 | 1 |
| Treatment should be made shorter | | 1 | 3 |
| REFUSAL AND HESITANCY TO GET SYPHILIS TREATMENT | | 4 | 9 |
| Hesitant | | 1 | 1 |
| Partners | | 1 | 2 |

| Name | Description | Files | References |
|---|---|---|---|
| Reasons for hesitancy | | 2 | 4 |
| Not hesitant | | 4 | 8 |
| ROLE | | 3 | 3 |
