## Supplemental File 6 for "*“Syphilis is a normal thing that children are born with…*”: Understanding Barriers and Facilitators to Maternal and Congenital Syphilis Care in Kampala, Uganda"

### COREQ (COnsolidated criteria for REporting Qualitative research) Checklist

*A 32-item checklist for interviews and focus groups*

**Instructions:** Enter the page number in the manuscript where each item is reported. If an item is not applicable or not reported, state this in the final column.

| No. | Topic | Guide questions/description | Reported on Page No. |
|---|---|---|---|
| **Domain 1: Research team and reflexivity** | | | |
| ***Personal Characteristics*** | | | |
| 1 | **Interviewer/facilitator** | Which author/s conducted the interview or focus group? | 19 |
| 2 | **Credentials** | What were the researcher's credentials? E.g. PhD, MD | Not Reported |
| 3 | **Occupation** | What was their occupation at the time of the study? | 19 |
| 4 | **Gender** | Was the researcher male or female? | N/A |
| 5 | **Experience and training** | What experience or training did the researcher have? | 6 |
| ***Relationship with participants*** | | | |
| 6 | **Relationship established** | Was a relationship established prior to study commencement? | 5 |
| 7 | **Participant knowledge of the interviewer** | What did the participants know about the researcher? E.g. personal goals, reasons for doing the research | 6 |
| 8 | **Interviewer characteristics** | What characteristics were reported about the interviewer/facilitator? E.g. bias, assumptions, reasons and interests in the research topic | 6 |
| **Domain 2: Study design** | | | |
| ***Theoretical framework*** | | | |
| 9 | **Methodological orientation and theory** | What methodological orientation was stated to underpin the study? E.g. grounded theory, discourse analysis, ethnography, phenomenology, content analysis | 5-6 |
| ***Participant selection*** | | | |

| No. | Topic | Guide questions/description | Reported on Page No. |
|---|---|---|---|
| 10 | **Sampling** | How were participants selected? E.g. purposive, convenience, consecutive, snowball | 5 |
| 11 | **Method of approach** | How were participants approached? E.g. face-to-face, telephone, mail, email | 5-6 |
| 12 | **Sample size** | How many participants were in the study? | 6 |
| 13 | **Non-participation** | How many people refused to participate or dropped out? Reasons? | 6 |
| ***Setting*** | | | |
| 14 | **Setting of data collection** | Where was the data collected? E.g. home, clinic, workplace | 5 |
| 15 | **Presence of non-participants** | Was anyone else present besides the participants and researchers? | 5 |
| 16 | **Description of sample** | What are the important characteristics of the sample? E.g. demographic data, date | 25-26 |
| ***Data collection*** | | | |
| 17 | **Interview guide** | Were questions, prompts, guides provided by the authors? Was it pilot tested? | 6 |
| 18 | **Repeat interviews** | Were repeat interviews carried out? If yes, how many? | Not done |
| 19 | **Audio/visual recording** | Did the research use audio or visual recording to collect the data? | 6 |
| 20 | **Field notes** | Were field notes made during and/or after the interview or focus group? | 6 |
| 21 | **Duration** | What was the duration of the interviews or focus group? | 6 |
| 22 | **Data saturation** | Was data saturation discussed? | 6 |
| 23 | **Transcripts returned** | Were transcripts returned to participants for comment and/or correction? | Not done |
| **Domain 3: Analysis and findings** | | | |
| ***Data analysis*** | | | |
| 24 | **Number of data coders** | How many data coders coded the data? | 6 |
| 25 | **Description of the coding tree** | Did authors provide a description of the coding tree? | 6-7 |
| 26 | **Derivation of themes** | Were themes identified in advance or derived from the data? | 6 |
| 27 | **Software** | What software, if applicable, was used to manage the data? | 6 |
| 28 | **Participant checking** | Did participants provide feedback on the findings? | Not done |

| No. | Topic | Guide questions/description | Reported on Page No. |
|---|---|---|---|
| *Reporting* | | | |
| 29 | **Quotations presented** | Were participant quotations presented to illustrate the themes/findings? Was each quotation identified? E.g. participant number | 7-14 |
| 30 | **Data and findings consistent** | Was there consistency between the data presented and the findings? | 7-14 |
| 31 | **Clarity of major themes** | Were major themes clearly presented in the findings? | 7-14 |
| 32 | **Clarity of minor themes** | Is there a description of diverse cases or discussion of minor themes? | 7-14 |

**Reference:** Tong A, Sainsbury P, Craig J. Consolidated criteria for reporting qualitative research (COREQ): a 32-item checklist for interviews and focus groups. International Journal for Quality in Health Care. 2007; 19(6): 349–357.
