## Supplemental File 3 for "*“Syphilis is a normal thing that children are born with…*”: Understanding Barriers and Facilitators to Maternal and Congenital Syphilis Care in Kampala, Uganda"

**Reducing maternal and congenital syphilis in Uganda; a mixed methods observational study.**

**Focus Group Discussion Guide**

**Introduction:**

Hi, my name is _____________ and I want to thank you for joining us today. I am working together with other scientists at the Infectious Diseases Institute and other institutions on this study in Kisenyi Health Centre IV, Kiswa Health Centre IV, Kawaala HC IV and Kawempe National Referral Hospital. We are conducting this research to explore your perceptions, attitudes and barriers to testing and treatment of maternal and congenital syphilis. I would like to say that there are no right or wrong answers in our conversation. We will simply be discussing your views, opinions and experiences on a range of topics. I would like to audio record our conversation as I may not be able to remember all your responses. Please do not be concerned about this: all measures will be taken by the researchers to maintain confidentiality of the interviews and discussions.

Information you tell us will **ONLY** be used for this research project. As we are audio recording the interview, we ask that you refrain from using names or identifying information of yourself or your partner. If at any time during the interview you feel uncomfortable you can ask for a break, refuse to answer any question, and are always free to leave. Do you have any questions before we start?

**Create rapport:** How have you been? How is work? How is the baby?

1. **Knowledge about syphilis**
   - Had you ever heard about syphilis before? (*Probe; how it is spread, signs and symptoms of syphilis, treatment of syphilis, effects of not treating syphilis to mother and child)*

2. **History of syphilis infection**
   - How did you come to know that you have syphilis? *(Probe whether its first time to test; history of prior syphilis diagnosis)*
   - How many months along were you when you found out that you have syphilis?
   - How did you feel when you found out that you have syphilis?

INFECTIOUS DISEASES INSTITUTE RESEARCH ETHICS COMMITTEE (IDI-REC) APPROVED VALID UNTIL 24 JUN 2023

3. **Treatment**
    - What are the perceived experiences in accessing the different services regarding syphilis
        - *Probe on how long it takes to test and get results*
        - *Probe on treatment – how long after testing does a patient receive treatment and how is the treatment*
        - *Probe on side effects – How they handle the side effects*

        - What would make a mother (or her partner) who has been diagnosed with syphilis refuse to get treatment

**2: Partner notification**

a) What would make it easy for someone to inform their partners that they are syphilis positive
b) What would prevent someone from telling the partner that they are syphilis positive
c) Why would you seek the help of a health worker or someone else to inform your partner about your syphilis results
d) How easy is it for men to seek professional syphilis related services?
e) What are the perceived benefits of informing your partner that you have syphilis?

**3: Birth outcomes**

a) What are your perceptions about having your child tested for syphilis at birth?
b) What are the perceived benefits of having a syphilis positive child treated early?
c) What are the perceived challenges of having a syphilis positive child not treated?

**General questions**

4. What do you like about the syphilis services that are offered here?



INFECTIOUS DISEASES INSTITUTE
RESEARCH ETHICS COMMITTEE (IDI-REC)
APPROVED
VALID UNTIL
24 JUN 2023

5. What don't you like about the syphilis services that are offered here?

6. What challenges does a person go through after finding out that they have syphilis? *Probe on partner notification, birth outcomes, treatment*

7. What are the perceived benefits from getting syphilis treatment?

8. Lessons learnt; what have you learnt while receiving syphilis related services?
9. Suggestions; Is there anything that you think should be changed or improved among the syphilis services offered at the health facility?

Thank you so much for taking time to participate in this study.

THE END

INFECTIOUS DISEASES INSTITUTE
RESEARCH ETHICS COMMITTEE (IDI-REC)
APPROVED
VALID UNTIL
24 JUN 2023
